# Investigating Environmental Determinants of Soil-Transmitted Helminths Transmission using GPS Tracking and Metagenomics Technologies

**DOI:** 10.1101/2023.07.17.23292808

**Authors:** Jeffrey Gabriel Sumboh, Kwasi Agyenkwa-Mawuli, Eyram Schwinger, Irene Owusu Donkor, Jewelna E. B. Akorli, Duah Dwomoh, Yvonne Ashong, Dickson Osabutey, Felix Owusu Ababio, Kwadwo Ansah Koram, Debbie Humphries, Michael Cappello, Samuel K. Kwofie, Michael D. Wilson

**Affiliations:** Department of Parasitology, Noguchi Memorial Institute for Medical Research, University of Ghana. Legon, Accra, Ghana; Department of Mathematics, University of Ghana. Legon, Accra, Ghana; Department of Epidemiology, Noguchi Memorial Institute for Medical Research, University of Ghana. Legon, Accra, Ghana; Department of Biostatistics, School of Public Health, University of Ghana. Legon, Accra, Ghana; Soil Research Institute, Council for Scientific and Industrial Research, Accra, Ghana; Department of Epidemiology of Microbial Diseases, Yale School of Public Health, New Haven, United States of America; School of Biomedical Engineering, Faculty of Engineering, University of Ghana. Legon, Accra, Ghana

**Keywords:** Soil-transmitted helminths, soil factors, GPS tracking, Metagenomics

## Abstract

**Background:** The Global Health community aims to eliminate soil-transmitted helminth (STH) infections by 2030. Current preventive methods such as Mass Drug Administration, WASH practices, and health education needs to be complimented to halt transmission. We tracked the movement of hookworm-infected and non-infected persons and investigated soil factors in the places they frequented within an endemic community to further understand the role of human movement and sources of infections.

**Methods:** 59 positive and negative participants wore GPS tracking devices for 10 consecutive days and their movement data captured in real time. The data was overlaid on the community map to determine where each group differentially spent most of their time. Soil samples were collected from these identified sites and other communal places. Physical and chemical properties were determined for each sample using standard methods and helminth eggs cultured into larvae using the Baermann technique. Bivariate and multivariate analyses were used to determine associations between larvae counts and soil factors. Helminth species were identified with metagenomic sequencing and their distributions mapped to sampling sites in the community.

**Results:** The study found that there was no significant difference in the average larvae counts in soil between sites assessed by infected and non-infected participants (*P*=0.59). However, soil factors, such as pH, carbon and sandy-loamy texture were associated with high larvae counts (*P*<0.001) while nitrogen and clay content were associated with low counts(*P*<0.001). The dominant helminth species identified were *Panagrolaimus superbus* (an anhydrobiotic helminth), *Parastrongyloides trichosuri* (a parasite of small mammals), *Trichuris trichuria* (whipworm), and *Ancylostoma caninum* (dog hookworm). Notably, no *Necator americanus* was identified in any soil sample.

**Conclusion:** This study provides important insights into the association between soil factors and soil-transmitted helminths. These findings contribute to our understanding of STH epidemiology and support evidence-based decision-making for elimination strategies.

**Author Summary:** Soil-transmitted helminth (STH) infections has been set to be eliminated 2030. To better understand how infections spread to achieve this, we tracked the movement of people positive and negative for infections. We enrolled 59 participants and equipped them with GPS tracking devices for 10 days continuously monitoring their movements in real time. We identified areas where each group spent the most time. Soil samples were collected from these locations and other communal areas. The physical and chemical properties of the soils were analysed using standard methods and helminth eggs cultured into larvae. Bivariate and multivariate analyses were used to study the relationship between larvae counts and soil factors. Metagenomic sequencing identified the types of helminths present in the soil samples. The study revealed that soil factors such as pH, carbon content, and sandy-loamy texture were associated with high larvae counts, while nitrogen and clay content were associated with lower counts. The dominant helminth species identified *were Panagrolaimus superbus, Parastrongyloides trichosuri, Trichuris trichiura* (whipworm), and *Ancylostoma caninum* (dog hookworm). Interestingly, no samples contained *Necator americanus*. This study provides important insights into the connection between soil factors and STHs infections enhancing our understanding of STH epidemiology to inform evidence-based strategies for eliminating.

## Introduction

Approximately 500 million individuals residing in tropical regions of Africa, South America, and Asia are infected with hookworms, predominantly *Necator americanus* and *Ancylostoma duodenale* [1]. These infections result in four million disability-adjusted life years (DALYs) [2] and incur an estimated cost of USD 139 billion in annual economic productivity losses [3] The primary clinical symptoms of hookworm disease include iron deficiency anaemia due to blood loss, abdominal pains, diarrhoea, and protein malnutrition [4]. In children and women of childbearing age, chronic infections and associated blood loss lead to low iron stores, which impairs physical and cognitive development in children and increases perinatal maternal/infant mortalities in pregnant women[5].

To combat hookworm infections, various treatment and control strategies have been implemented, including Mass Drug Administration (MDA), sanitation improvements and health education programs[6]. These strategies have been effective in reducing the prevalence of hookworm infection, but there is still a need for further research and implementation to completely eradicate this disease. The high rates of reinfection following drug treatment and emerging drug resistance pose major challenges to current deworming programs[7]. Hookworm infections are mainly treated with single doses of either albendazole (400 mg) or mebendazole (500 mg)[4], [8] termed preventive chemotherapy (PC) and is based on MDA to all at-risk individuals in endemic areas[9], [10]. As of 2016, 638.5 million people had been covered, which included 69.5% of at-risk school-aged children and 50.8% of at-risk pre-school-aged children globally [11], [12]. These efforts were further supplemented with the implementation of the Water, Sanitation and Hygiene (WASH) programs to address the lack of potable water and sanitation facilities in schools[13].

The situation in Ghana is comparable to other countries where parasitic infections, specifically hookworms, are prevalent, with reported rates as high as 50% [11], [14], [15]. Our cohort studies conducted in rural communities in the Kintampo North Municipality (KNM), Ghana have shown that prevalence reduces from around 40% to 5% and has remained at this level after 5 cycles of biannual treatment with albendazole[16], [17]. Hookworm infection is more widespread in resource-poor rural communities situated in the deciduous forest belt. Unfortunately, these areas are often also affected by other endemic infections such as schistosomiasis, malaria, HIV, and tuberculosis [18]–[22] The coexistence of these diseases exacerbates the negative impact of hookworm and other parasitic infections [17], [23], [24], highlighting the need for targeted efforts to address the disease transmission dynamics.

Ghana has joined the Global Health effort to eliminate Soil-Transmitted Helminths (STHs) by 2030 [25]. The plan to achieve the set goal continues to be mass administration of benzimidazoles, along with health education and Water, Sanitation and Hygiene (WASH) strategies. However, a critical understanding of the factors that influence persisting infections must be addressed to ensure success[12]. The prevalence and distribution of hookworm infections are associated with several spatial and temporal factors, including space, time, seasons, and socio-economic status[26], [27]. Specific geographical locations, temperature and humidity facilitate the growth and development of larvae in the soil. Warm and humid weather conditions create suitable conditions for the eggs to thrive[28]. Socio-economic indices i.e., poverty, lack of education, and inadequate sanitation facilities increase the risk of infections and limit access to health care and hygiene education, which are essential for preventing these infections[29]. Inadequate toilet facilities and open defecation lead to faecal contamination of soil, promoting the spread of hookworms[30].

Recent health trends and services are increasingly influenced by the built environment and the use of Global Positioning Systems (GPS) has gained popularity, as it provides an accurate means of locating phenomena concerning important disease epidemiological variables. GPS technology has become a valuable tool for health researchers, particularly in studying geographically dependent diseases, such as hookworm [31].

We, therefore, employed GPS technology to monitor the movements of hookworm-infected and non-infected participants to identify the sites in the community that were likely sources of infections and the soil properties that are associated with the presence of helminth larvae.

## Materials and Methods

### Study area

Kawampe, the study site is a rural community in the Kintampo North Municipality (KNM), which is located between latitudes 8°45’N and 7°45’N and longitudes 1°20’W and 2°1’E and, covers an area of 5108 square kilometers in the forest-savanna transition zone of the middle belt of Ghana (Figure 1). The soil type at KNM is classified as predominantly forest ochrosols, which have high percentages of magnesium, calcium and lime making the soil less acidic and more alkaline[32]. Kawampe has a population of approximately 3000 inhabitants, 85% of them belonging to the Kokomba ethnic group and predominantly Christians. The vegetation is woody savanna grassland interspersed with forest covers [33]. The main occupations are farming and charcoal production[34]. The area experiences two rainy seasons, the major and minor from May to July and September to October, respectively[35], [36]. The hookworm prevalence at Kawampe has dropped significantly following baseline studies there in 2015 [34], [37]. It is currently at 9.2% in 2018 and 5.3 % in 2020 (Wilson et. al, *unpublished*).

**Figure 1:**
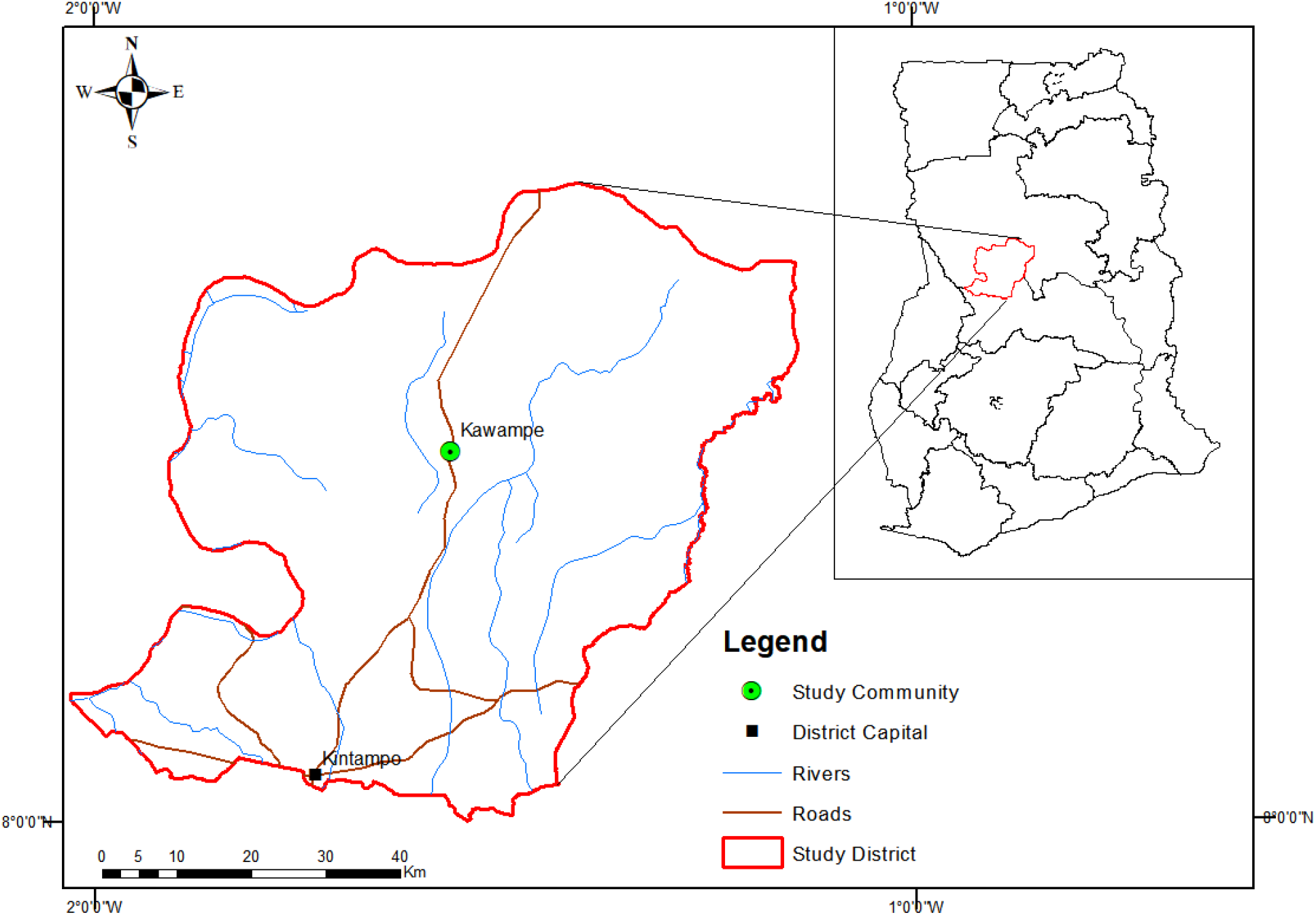
Map of Ghana showing the Kintampo North Municipal area. Kawampe is located on the main trunk road to the north of Ghana. *Map created with ArcGIS 10.8 with shapefiles from* https://www.diva-gis.org/gdata.

### GPS tracking of hookworm-infected and non-infected cases

Participants aged 6 years and above were screened for STH infections using Kato-Katz [21], [22]. Fifty-nine participants comprising 30 adults and 29 children were randomly assigned to positive and negative infection groups. They were subdivided into six (6) groups of 9-10 each. At any point in time, 20 participants were tagged with GPS tracking devices (i-gotU and Globalsat DG-100 data logger) and followed for 10 continuous days. The GPS devices recorded coordinates every 6 to 10 seconds allowing for the movement patterns of each participant to be plotted. The data was downloaded to a dedicated laptop using the device’s software and exported to ArcGIS 10.8 and Python 3.9 to produce a 20-by-20-meter grid map of the community. The K-means obtained from the analyses were used to determine the centre point of each grid. The exact coordinates of each individual in a group’s movement, the time spent at a particular area and how far they travelled were determined and overlaid on the grid map. The sites that were differentially visited most by the two groups were identified. Soil samples were taken from the central point of each visited grid, and public places of interest such as rubbish dumps, public toilet facilities, water sources, children’s playgrounds, schools and places of religious activities[38].

### Soil sampling

The Auger soil sampler was used to collect the soil from the identified sites and communal places of interest[39]. The soil was scooped from a 5cm depth, placed in a container and mixed by shaking vigorously. Each soil sample was then divided into two Ziplock bags and transported to the laboratories. One part was used for culturing helminth eggs to obtain larvae and, the other for the determination of physical and chemical properties.

### Soil physical and chemical analyses

A total of 21 parameters were measured: soil pH, electrical conductivity, total nitrogen, organic carbon, available phosphorus, Effective Cation Exchange Capacity (ECEC), Exchangeable Acidity and Hydrogen, micro-nutrients (Fe, Cu, Mn and Zn), clay and sand content, and soil texture. The soil pH was measured in the supernatant of a 1:2.5 soil-water ratio using a glass electrode pH meter with an H19017 Microprocessor [40]. Total nitrogen was determined by the Kjeldahl digestion and distillation procedure[41]. The soil organic carbon was determined by the modified dichromate oxidation method of Walkley-Black[42]. The readily acid-soluble forms of phosphorus were extracted with HCl: NH4F mixture (Bray’s No. 1 extract) and determined calorimetrically by ascorbic reduction[43], [44].

Effective cation exchange capacity (ECEC) was determined by the sum of exchangeable bases (Ca^2+^, Mg^2+^, K^+^ and Na^+^) and exchangeable acidity (Al^3+^ + H^+^). These exchangeable bases were determined in 1.0M ammonium acetate (NH_4_OAc). The calcium and magnesium were determined by EDTA titration and, potassium and sodium were determined by flame photometry [45]. The Exchangeable acidity (Al^+^+H^+^) was determined in 1.0M KCl extract. The soil micro-nutrients (iron, copper, manganese, and zinc) were extracted by ethylenediamine tetra acetic acid (EDTA) with ammonium acetate method and measured with an Atomic Absorption Spectrophotometer [46].

### Culture of helminth eggs

The Baermann technique was used to culture helminth eggs in the soil into larvae. Approximately 475g of soil was weighed and dampened thoroughly with tap water. The damp samples were then transferred onto Kimwipe paper tissue wrapped neatly, placed onto a standardized mesh and placed gently in the funnel of the Baermann setup with the clamp of the tubbing closed. Lukewarm water at about 37°C was poured gently onto the funnel until the wrapped soil samples were completely immersed. It was then allowed to stand for 18 hours after which 200-300mL of water was collected from the bottom of the set-up by slowly releasing the clamp. The collected water was observed under the microscope for the presence of larvae. The number of larvae harvested from each soil sample was estimated and stored in RNALater at −20°C.

The larvae were thawed at room temperature and washed in PBS, pulse vortexed and 10µL of the solution was transferred onto a petri dish and viewed under the microscope for confirmation of the presence of larvae. Genomic DNA was extracted from approximately 10 larvae of each sample using the DNeasy® Blood and Tissue kit (Cat. No. 69506, QIAGEN,). Before tissue lysis with ATL, 100µL of larval suspension was frozen at −80 C for an hour and macerated. The extraction process followed the manufacturer’s instructions thereafter and, DNA was eluted in 100μL of elution buffer. The concentration and purity of the isolated DNA were determined using a BioDrop ND-2000 spectrophotometer (NanoDrop, Wilmington, DE, USA). Two no-template control extractions were also included in the process to control for contamination. The extracted DNA was stored at −20°C until further use.

### Metagenomics sequence analyses

A total of 40 DNA samples were submitted for shotgun metagenomic sequencing; 36 passed initial sample quality control checks and were used in further library preparation and sequenced. 1,270,226,294 (190.5G) raw reads were obtained with an average of 42,340,876 per sample. Read quality checks were performed on all 36 samples using the *FastQC* [47] and *MultiQC* [31]. Low-quality reads and adapters were trimmed off with *Cutadapt* [48] and quality checks were repeated after trimming.

The trimmed reads for each sample were run against a database with assigned taxonomy information using *Kraken2* [49], which is a taxonomic classification system and database that uses exact k-mer matches to achieve high accuracy and fast classification speeds. There are no available standard *Kraken2* taxonomy databases built for helminths, therefore, a customized database named *NematodeDB* was built from the genome sequences of about 160 flatworm and nematode species. The taxonomic information of the genomes was obtained from the *NCBI* reference sequences database[50]. The processed reads were then run against *NematodeDB* to yield *Kraken2* reports on the mapped or classified reads and their taxonomy assignments.

### Visualization

The *Kraken2* reports for all the samples were compressed and analysed using *Pavian* [51]. A Sankey diagram for each sample was generated as well as the summaries of taxonomy assignments of the reads. Since a customized database for only worms was used, the worm hits were classified under the *Microbial reads* column in *Pavian* while the columns for other organisms remained empty. Two *.xlsx* files were then generated from *Pavian* as follows: *Identified species and abundances* containing 100 species of worms identified and the number of mapped (classified) reads across the samples, *Classification summary* containing the numbers and percentages for both classified and unclassified reads. The control sample also had some hits, which were inferred as artefacts. This served as a guide for examining the same species for the other samples, which were considered if their abundance were above the ones reported in the control.

### Ethical Considerations

The study proposal was approved by the Noguchi Memorial Institute for Medical Research Institutional Review Board (NIRB #100/16-17), the Kintampo Health Research Center Ethics Review Committee (#KHRCIEC 2017-20) and NIH/NIAID (DMID Protocol #17-0061). Permission was obtained from the chief, elders, and opinion leaders of the study community. Informed consent was obtained from adults and assent/parental consent in the case of children.

### Data analyses

To determine which soil parameters were significantly associated with the larval counts we employed the Negative Binomial Regression Model (NBRM) method to determine the associations between the soil factors and larval counts. The NBRM provides flexibility in handling overdispersion and accommodating different relationships[52] between the soil factors and the (larval counts).

## RESULTS

### The study participants

Out of the 59 participants who were monitored, 54.2% (n=32) were males, and 45.8% (n=27) were females. The average age was 25.9 years (SD=±18.6; Range=6-83), and the majority (33.9%; n=20) were aged between 11 and 20 years. Hookworm was present in 52% of the participants. Both children and adults constituted equal proportions of both positive and negative participants.

### Soil characteristics of the study site

Two types of soil; the Lima series (Eutric Planosol) and Kumayili series (Plinthic Lixisol)[53] existed at Kawampe. The Lima series consisted of 30-60cm of light brownish-pinkish grey fine sandy loam or loamy fine sand, over a thin layer (15-20cm) of very pale brown or pinkish grey fine sandy loam containing frequently polished ironstone concretions. This immediately covered grey silty clay that is very hard and very compact and develops into a clay pan when dry and into a plastic and massive pan when wet. This layer extends to over 120cm from the surface and may occasionally have polished ironstone concretions in the top part of this layer.

The Kumayili series occurred on low summits, and upper and middle slope sites with a 2-3 % gradient. The topsoil consisted of about 15 cm to 30 cm of dark brown to reddish-brown sandy clay loam or sandy loam over a yellowish-red sandy loam or sandy clay loam to a depth of 100-125 cm. Beneath this layer is a tightly packed either ironstone concretions or iron pans.

The Lima soil type was found mainly to the north of Kawampe at 10 sampled sites and, the rest of the sites were Kumayili (Figure 2).

**Figure 2:**
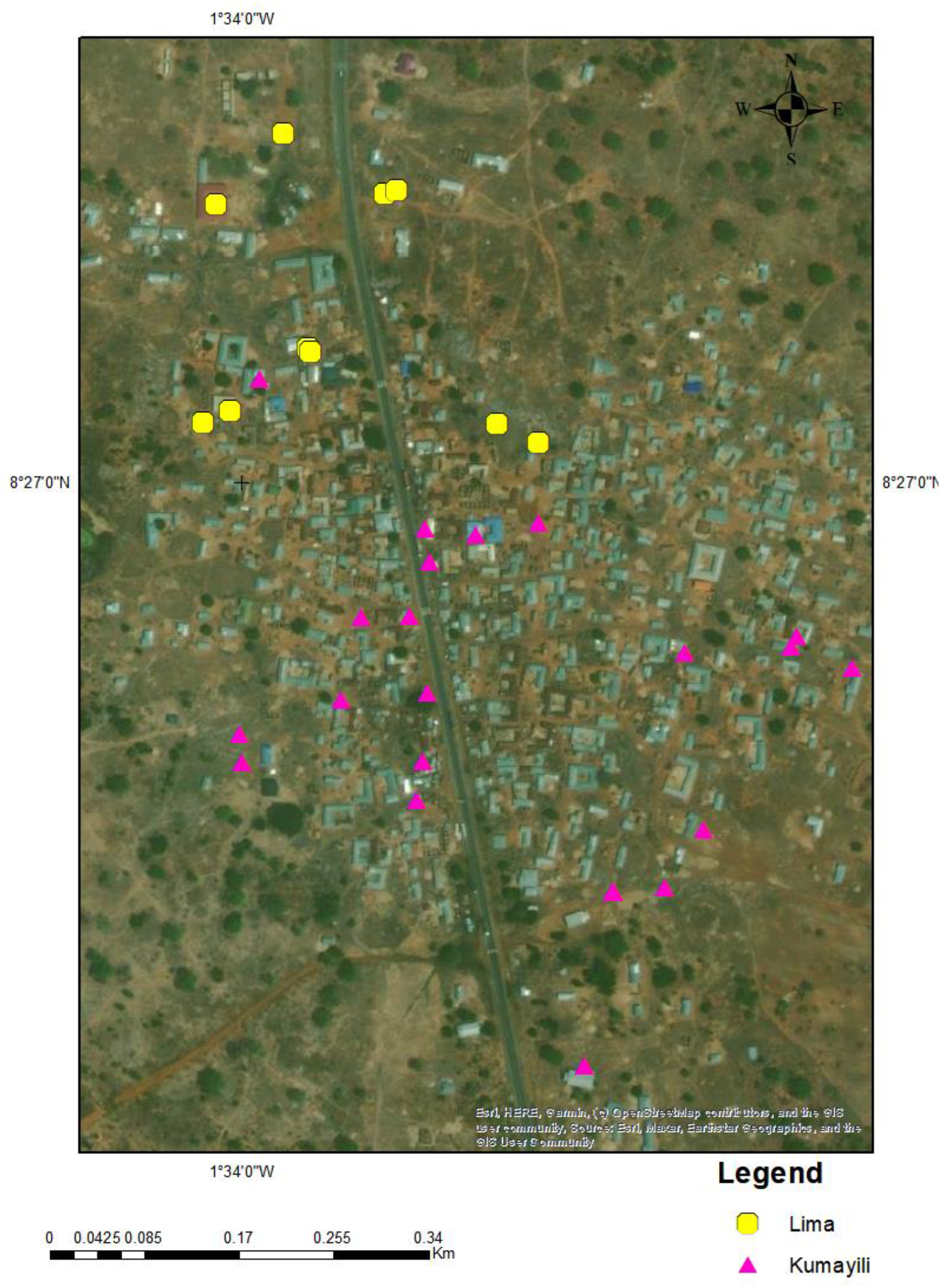
Map of Kawampe showing the soil types at each sampled site. Lima series (yellow dots) was found mainly to the north of the community while the Kumayili type (pink triangles) was widespread and to the southern part. *Map created with ArcGIS 10.8*

### Spatial movements of the study participants

The participants’ daily movements were mainly within the community and to neighbouring farmlands (Figure 3). These movements were mostly in the mornings and evenings and virtually non-existent at night. The movement dynamics of both positive and negative adult participants indicated a close interaction among community members (Figures 3A and 3B). Their daily movements became more individualized due to the distinct ownership of farms (Figure 3B). The movements of children primarily revolved around the community, as their activities were centred on community and school-based engagements.

**Figure 3:**
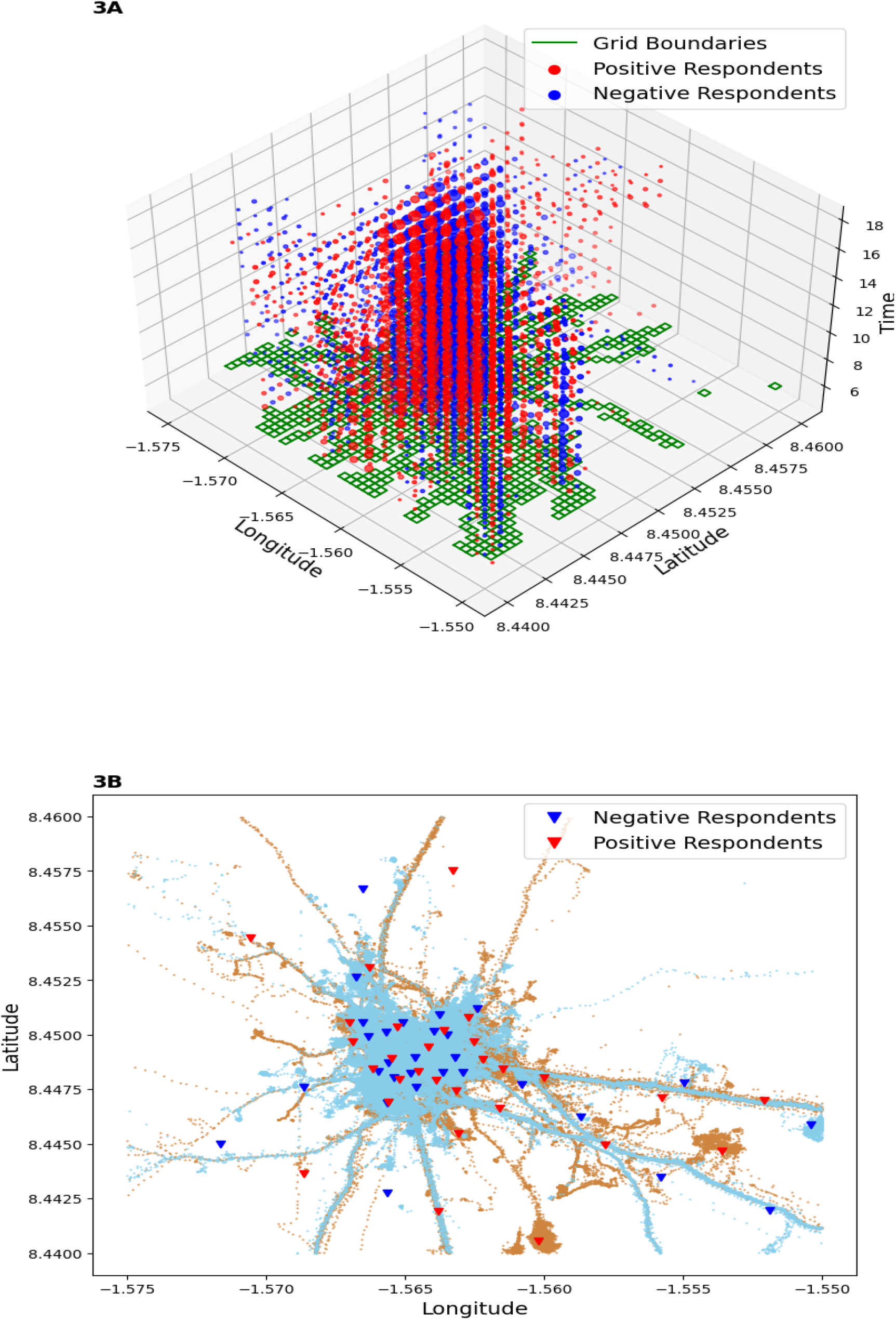
3A) 3-D illustration of all the participants’ movements across space and time. The green patches were the areas where most participants were located, while the red and blue circles indicated the length of time both positive and negative participants spent within these green patches respectively. The 3B) 2-D plot illustrates the movement patterns of the positive group (red lines) and negative group (blue lines). Red triangles denote the sites frequented by infected individuals and where soil samples were collected, while blue triangles represent where negatives were mostly found. These triangles indicate the exact centers of the grids (green patches) in 3A.

### Association of soil factors with helminth larval density

At least one larva was found at every site where soil samples were collected. The highest number of larvae (n=193) was observed in soil from a church area with a sandy loam texture and pH of 5.765. The second highest number of larvae (n=165) was detected in soil from the marketplace, which had a pH of 6.84 and a loamy sand texture.

Soil factors that produced high larvae counts included soil pH, sandy-loamy soil type, and effective cation exchange capacity (Figure 4). An increase in soil pH level was associated with an incidence rate ratio of 3.7, (CI; 2.1-6.6, *P<* 0.001) based on the larvae counts and soil parameters from the NBRM. Sandy-loamy soil had approximately 7 times more larvae compared to loamy soil, with an incidence rate ratio of 6.7, (2.3-19.7, *P<*0.001) and Effective cation exchange capacity had an incidence rate ratio of 1.3, (1.2-1.43, *P*<0.001).

**Figure 4:**
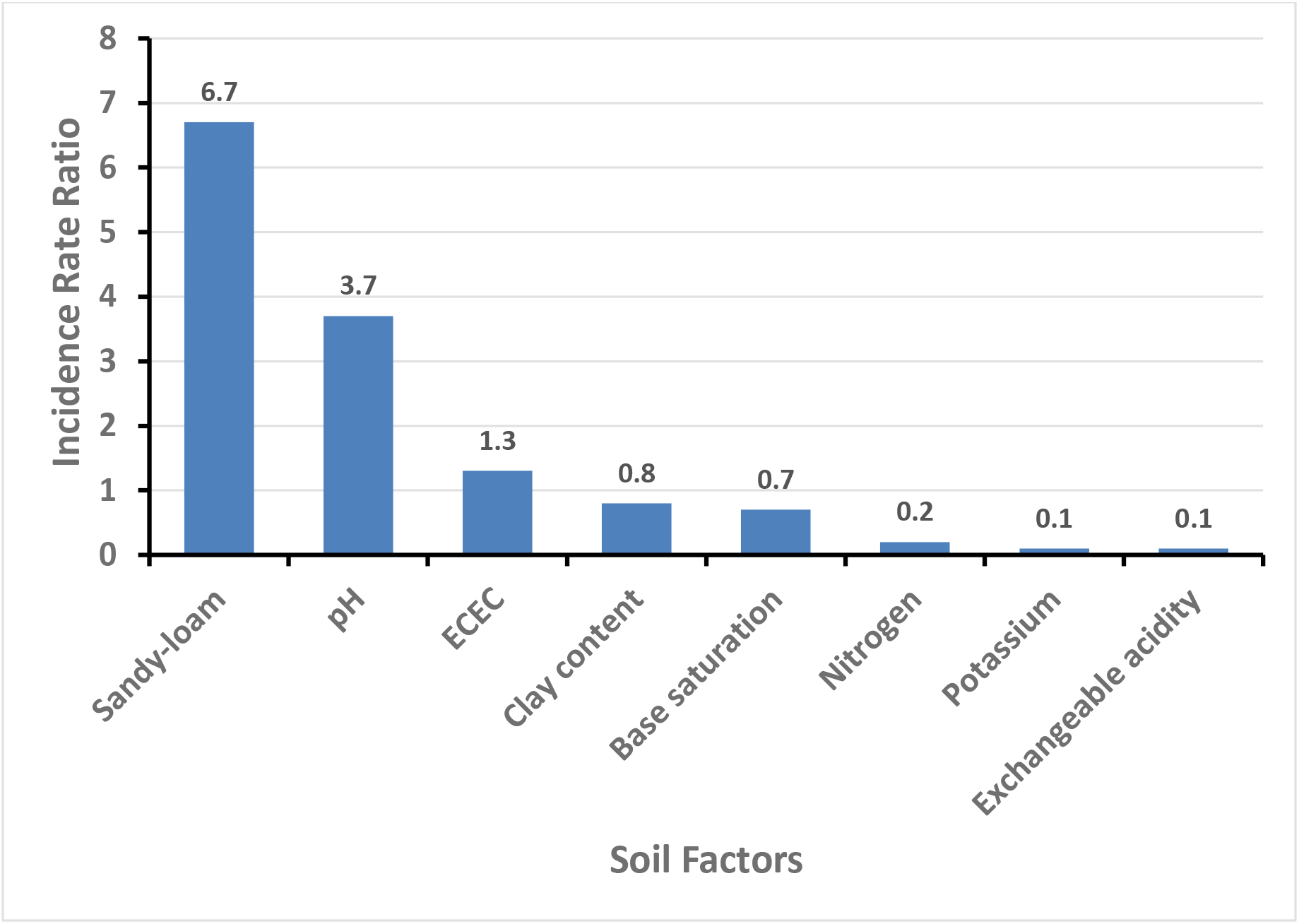
Soil factors significantly associated with high and low larvae counts. High levels of potassium, exchangeable acidity, base saturation, clay, and nitrogen content on the other hand were associated with low larvae counts (Table 4). Potassium content had an incidence rate ratio (IRR) of 0.1, (0.1-0.3*, P<*0.001). Exchangeable acidity had an IRR of 0.01, (0.00-0.3, *P=*0.002) and that of the base saturation was 0.7, (0.4-0.9, *P=*0.042). High clay content had an IRR of 0.8, a confidence interval of 0.7-0.9, and a p-value of less than 0.001. High nitrogen content had an IRR of 0.2, (0.1-0.4, *P<* 0.001), which was above the median value.

### Identification of helminth species

Four dominant species were identified in the soil samples: *Parastrongyloides trichosuri, Panagrolaimus superbus, Trichuris trichiura* and *Ancylostoma caninum* (Table 1, Figure 5). Interestingly, there were no hits for *Necator americanus* (human hookworm).

**Figure 5:**
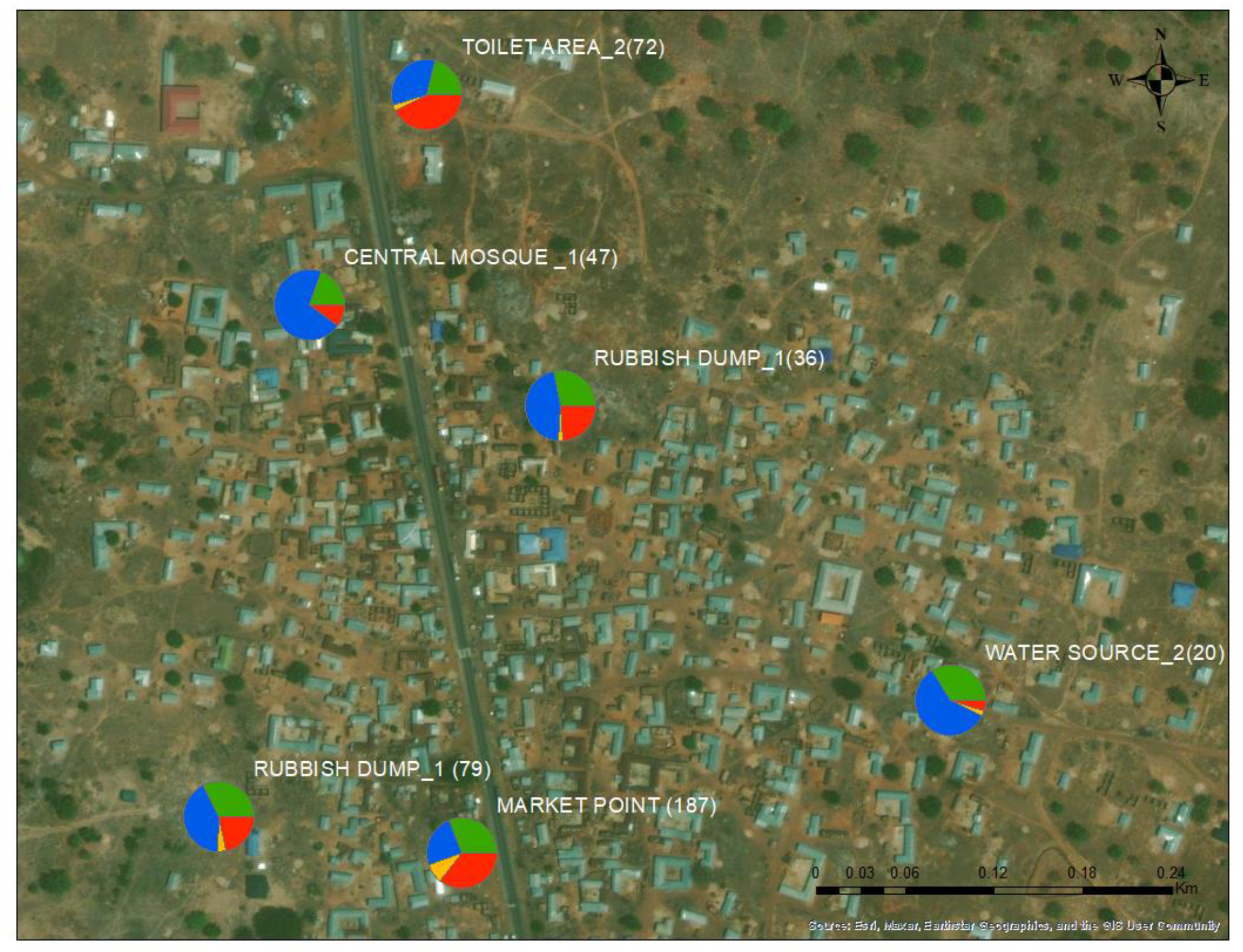
The distribution of dominant helminth parasites found in specific areas within the study community. *Parastrongyloides trichosuri* (blue), *Panagrolaimus superbus* (green, *Trichuris trichiura* (yellow) and *Ancylostoma caninum* (red). The counts are in parentheses.

**Table 1:** The location and distribution of the main helminth species cultured from the soil samples and identified using shotgun metagenomics.

| Location | Larvae<br>Count | *P. trichosuri*<br>(%) | *P. superbus*<br>(%) | *T. trichiura*<br>(%) | *A. caninum*<br>(%) |
| --- | --- | --- | --- | --- | --- |
| Market point | 187 | 11.55 | 14.6 | 4.2 | 16.85 |
| Rubbish dump_1 | 79 | 13.2 | 10.15 | 1.05 | 6.75 |
| Central mosque_1 | 47 | 42.15 | 11.75 | 0.45 | 6.25 |
| Toilet area_2 | 72 | 11.5 | 7.2 | 1 | 14.5 |
| Rubbish dump_2 | 36 | 16.8 | 10.1 | 0.9 | 8.6 |
| Water source_2 | 20 | 17.8 | 10.1 | 0.8 | 1.4 |

## Discussion and Conclusion

The global health community is targeting 2030 as the year to achieve the elimination of STH. Thus, any additional intervention tool that could support the existing MDA and WASH strategies is a prerequisite for this to be achieved. Our studies, therefore, focused on tracking hookworm-infected individuals and gathering soil characteristics to identify specific areas in a community that may contribute significantly to hookworm transmission[54]. Based on our study, the pH level of soil was found to be a critical determinant of parasite growth, as it was linked to elevated larval counts. The influence of soil pH on parasite development is connected to the soil’s acid-base equilibrium. Parasites that undergo development in soils with elevated pH levels acquire enhanced tolerance towards alkaline conditions. This heightened tolerance enables them to flourish and reproduce with greater efficiency. Conversely, the parasites that developed in soils with lower pH levels become more susceptible to alkaline conditions, impeding their growth and reproductive capabilities [55]. The levels of exchangeable cations and exchangeable acidity (ECEC) primarily depend on the presence of sodium, potassium, calcium, and magnesium in the soil, and are known to facilitate the absorption of nutrients essential for the survival of flora and fauna[56]. Zinc is another crucial factor that contributed to elevated larvae counts. It is a vital mineral for soil-borne parasites, forming a critical component of enzymes, proteins, and DNA. Incorporating zinc into the soil promotes the growth of parasitic nematodes and mitigates damage caused by pests[57].

In this study, the soil type that exhibited the least conducive environment for larval density was clay. Clay soil has limited air circulation and pore spaces, making it unsuitable for larvae habitation. Soils that were primarily clay-based were found to have lower larvae counts[58]. Other factors that had a significantly low impact on larval density included carbon content, base saturation, and nitrogen levels in the soil. Plants and nematode growth have a symbiotic relationship, flourishing on cultivated lands with aggregate quantities [32]. Carbon, base saturation and nitrogen improve growth in such cases. The soils sampled were on uncultivated lands hence the low larvae count[59].

The predominant parasite discovered in all soil samples was *Panagrolaimus superbus*, an anhydrobiotic nematode capable of surviving extreme environmental conditions such as water scarcity, high pressure, and temperature variations[60], [61]. This is the first report of this species in Ghana. *Panagrolaimus* nematodes have colonized various habitats, including arctic and antarctic biomes, as well as arid deserts. Most of these species are capable of cryptobiosis, and many are parthenogenetic, allowing them to survive repeated desiccation and freezing cycles. The medical significance of *P. superbus* in humans remains unknown. *Parastrongyloides trichosuri* was the second most abundant helminth species found, and its discovery in Ghana was unexpected as it is a nematode parasite native to Australasia, commonly infecting small mammals such as the Australian Brush-tailed possums [62]. There is a high population of small mammals in Ghana, particularly rodents, which can be hosts for parasites commonly found around rubbish dumps. *P. trichosuri* may have been introduced into the soil from such rodents. *P. trichosuri* is an interesting nematode that can undergo multiple reproductive cycles as a free-living worm, enabling it to increase the number of its infective L3s. Thus, it can be maintained in cultured conditions for several free-living life cycles without requiring a permissive host, usually its marsupial host. *Trichuris trichiura*, also known as the human whipworm, was the third most common soil-transmitted helminth (STH) causing trichuriasis [63]. In severe infections, trichuriasis causes frequent, painful bowel movements, rectal prolapse, and stunted growth in children. Although only minimally present in 21 out of the 32 soil samples, it was abundant in the remaining samples with high read counts. *Ancylostoma caninum*, commonly known as the dog hookworm, is a parasitic nematode that infects dogs and cats, causing eosinophilic enteritis in humans[64]. This species was found in all samples, albeit in lower numbers compared to the other species mentioned above. Cutaneous Larva Migrans (CLM), a condition caused by some species of hookworms, is associated with this species. Larvae enter humans through direct contact with the skin, commonly transmitted through faeces deposited in the soil.

The presence of parasites was observed in varying proportions in the two soil types (Kumayili and Lima) found in the community. *Parastrongyloides trichosuri* was the most prevalent parasite in all the soil samples (>50% prevalence), suggesting an abundance of small mammals in the community. Kumayili soil had a higher prevalence of *Ancylostoma caninum* and *Panagrolaimus superbus* compared to Lima soil. *Trichuris trichiura*, the only parasite identified in this study that infects humans, was found to be twice as prevalent around the market area and associated with Kumayili soil. Human hookworm species, *Necator Americanus* and *Ancylostoma duodenale* were notably absent in the soils sampled, although they are known to usually thrive in warm and moist environments with sandy loamy soils and sufficient organic matter content similar to the conditions in Kawampe [65]. Several potential reasons could account for this absence. First, the soil samples were collected at the end of the rainy season. It is important to note that regions characterized by heavy rainfall and warm temperatures are often associated with high transmission rates of Necator americanus[66], Second, it is possible that the soil sampling did not encompass areas within the community where the parasites are more likely to be present[67]. Also, the time of sampling might have influenced the larval load, resulting in low detection rates during the sampling period[68]. Lastly, the parasites may be localized to specific locations within the community which suggests that a more elaborate sampling needs to be done to increase the chances of finding them [69], [70].

This study on soil factors for soil-transmitted helminths (STH) has provided valuable insights into the relationship between soil characteristics and the presence of some STH larvae. Tracking the movements of participants has also shown significant convergence of people, whether infected or not in particular places within the community. Sanitation and proper waste management in such concentrated areas may reduce soil infestation.

## Data Availability

All data generated or analyzed during this study are included in this published article.

## Authors’ Contributions

MDW and MC conceived the project. JGS collected the field data, analysed and drafted the manuscript, ES analysed the GPS movement data, KAM performed the metagenomics analysis, DD, IOD, JA, KAK, DH, SKK and YA supervised the study, FOA and DO performed the soil analysis, and all the authors reviewed the manuscript and approved it.

## Acknowledgements

We appreciate the support of the NIINE laboratory and fieldwork teams, along with that provided by the staff of the Kintampo Health and Research Centre. We are also grateful to the chiefs, community leaders and participants in the Kintampo-North Municipality.

